# SARS-CoV-2 variants in Paraguay: Detection and surveillance with a readily modifiable, multiplex real-time RT-PCR

**DOI:** 10.1101/2021.09.15.21263618

**Authors:** Magaly Martinez, Phuong-Vi Nguyen, Maxwell Su, Fátima Cardozo, Adriana Valenzuela, Laura Franco, María Eugenia Galeano, Leticia Elizabeth Rojas, Chyntia Carolina Díaz Acosta, Jonás Fernández, Joel Ortiz, Florencia del Puerto, Laura Mendoza, Eva Nara, Alejandra Rojas, Jesse J. Waggoner

## Abstract

**Objectives:** The objective of the current study was to develop a lower-cost and scalable protocol to identify and monitor SARS-CoV-2 variants in Paraguay by pairing real-time RT-PCR detection of spike mutations with amplicon Sanger sequencing and whole-genome Nanopore sequencing.

**Methods:** 201 acute-phase nasopharyngeal samples from SARS-CoV-2-positive individuals were tested with two rRT-PCRs: 1) N2RP assay to confirm SARS-CoV-2 RNA detection (CDC N2 target), and 2) the Spike SNP assay to detect mutations in the *spike* receptor binding domain. The assay was performed with probes to identify mutations associated with the following variants: alpha (501Y), beta/gamma (417variant/484K/501Y), delta (452R/478K), and lambda (452Q/490S).

**Results:** All samples were positive for SARS-CoV-2 in the N2RP assay (mean Ct, 20.8; SD 5.6); 198/201 (98.5%) tested positive in the Spike SNP assay. The most common genotype was 417variant/484K/501Y, detected in 102/198 samples (51.5%) and most consistent with P.1 lineage (gamma variant) in Paraguay. No mutations (K417 only) were found in 64/198 (32.3%); and K417/484K was identified in 22/198 (11.1%), consistent with P.2 (zeta). Seven samples (3.5%) tested positive for 452R without 478K, and one sample with genotype K417/501Y was confirmed as B.1.1.7 (alpha). Results were confirmed by Sanger sequencing in 181/181 samples (100%) with high-quality amplicon sequences, and variant calls were consistent with Nanopore sequencing in 29/29 samples.

**Conclusions:** The Spike SNP assay provides accurate detection of mutations associated with SARS-CoV-2 variants. This can be implemented in laboratories performing rRT-PCR to improve population-level surveillance for these mutations and inform the judicious use of scarce sequencing resources.

## Introduction

Amidst the global pandemic caused by severe acute respiratory syndrome coronavirus 2 (SARS-CoV-2) numerous variants have emerged due to mutations in the positive-sense RNA genome. Variants of concern (VOCs) and variants of interest (VOIs) bear mutations that impact detection, treatment, clinical severity, transmission, and/or immune protection from prior infection or vaccination [1]. Whole genome sequencing is the reference method for detecting and tracking variants, but this is a time- and resource-intensive process with capacity varying markedly between regions [2-5]. It has been estimated that 5% of all samples need to be sequenced to reliably detect variants at a prevalence of 0.1-1% [3], yet for many regions, this goal remains unattainable. Limited available sequencing capacity and overwhelming case numbers have motivated the development of real-time reverse transcriptase PCRs (rRT-PCRs) for the detection and surveillance of specific mutations found among VOCs/VOIs [6-20]. For these reasons, our group designed the Spike SNP assay, which is multiplex a rRT-PCR designed to detect specific mutations in the receptor binding motif of the Spike receptor binding domain (RBD) (Figure 1) [20].

**Figure 1.**
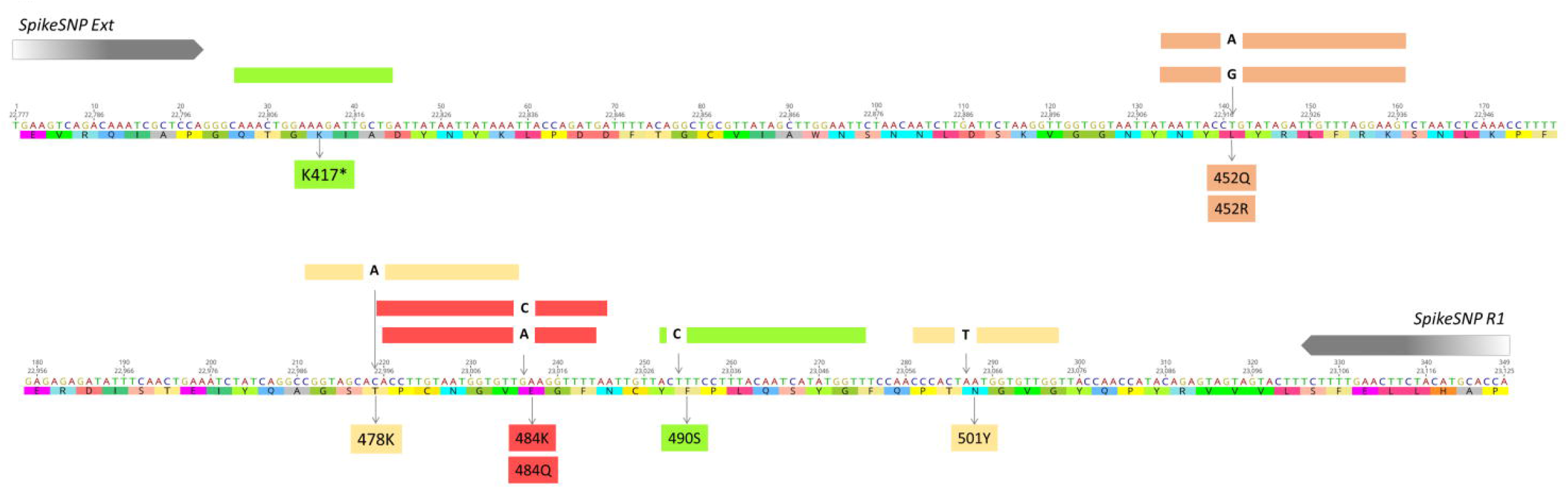
Graphical display of the 348-base-pair Spike SNP target including the nucleic acid sequence and amino acid translation of the Wuhan-Hu-1 strain (NC_045512.2). The locations of the forward and reverse primer sequences and all hydrolysis probes are shown. Probe colors correspond to fluorophores on each probe and the detection channel in the Rotor-Gene Q instrument. Detected nucleotide changes are displayed within the probe, and amino acid mutations are shown below the sequence. The probe for K417 is marked (*) as this is the only probe for which variant sequences result in signal loss and this has only been successfully designed with LNA bases.

In Paraguay, the first confirmed SARS-CoV-2 infection was reported on 07 March 2020 [21]. As of 25 August 2021, 457,971 cases were reported in the country with 228 whole genome sequences (0.05% of cases) available in the GISAID database (www.gisaid.org) [22]. Published reports on case series in Paraguay have been limited to descriptive analyses of symptoms and disease outcomes [23, 24]. As such, little is known about variants that circulated in the country during the beginning of the pandemic in 2020. However, data from 2021 show that variants originally detected in Brazil, such as the P.1 (gamma variant) and P.2 (zeta) lineages have circulated widely (GISAID database). Nanopore sequencing on the portable MinION sequencer has recently become available in Asunción, the capital of Paraguay, to detect and monitor SARS-CoV-2 variants for the first time, and this technology provides lower cost and more rapid whole genome sequencing compared to other methods [2, 25]. The objective of the current study was to develop an economical and scalable protocol based on the Spike SNP assay and Nanopore sequencing to identify and monitor VOCs/VOIs in acute-phase samples in Paraguay.

## Methods

### Clinical samples

Nasopharyngeal (NP) samples were obtained from residual baseline surveillance samples at the Instituto de Investigaciones en Ciencias de la Salud, Universidad Nacional de Asunción (IICS-UNA) Laboratory. Samples were deidentified, aliquoted and stored at -80°C until nucleic acid extraction. A convenience set of 201 SARS-CoV-2 positive samples was chosen for this study. Samples tested positive for the envelope gene target of the Charité protocol [26] with a cycle threshold (Ct) <34 and were collected from different cities in Paraguay from symptomatic and asymptomatic individuals. This study was reviewed and approved by the IICS Scientific and Ethics Committee (P38/2020) and the Emory Institutional Review Board (study 00110736).

### Spike SNP design and performance

Hydrolysis probes were designed to detect mutations associated with emerging variants of concern: L452Q (CAG>CTG) and F490S (UUU>UCU, C.37, lambda), E484Q (GAA>CAA, B.1.617.1, kappa), and T478K (ACA>AAA, B.1.617.2, delta). All new probes were designed as described [20], without locked nucleic acids (LNA). Unmodified probes for 484K and 501Y were also designed and evaluated. New probes were individually evaluated using SARS-CoV-2 genomic RNA in singleplex assays. Probes that provided the most sensitive signal based on Ct value, with preserved specificity, were selected. Subsequently, probes were tested in combination and compared side-by-side with the corresponding singleplex assays. Probe evaluations were performed utilizing characterized variant control samples kindly provided by the National Institutes of Health Variant Task Force.

Nucleic acids were extracted from all samples on an EMAG instrument (bioMérieux, Durham, NC) from 200µL of NP swab and eluted in 50µL. All rRT-PCRs were performed on a Rotor-Gene Q instrument (Qiagen, Germantown, MD) using 20µL reactions of the Luna Probe One-Step RT-qPCR Kit (New England Biolabs, Ipswich, MA) and 5µL of nucleic acid eluate. Each sample was tested in a duplex reaction for the SARS-CoV-2 nucleocapsid 2 target and RNase P (N2RP) as well as two different reactions of the Spike SNP assay including probes for 1) K417, 452R, 484K, and 501Y, as described [20]; and 2) 452Q, 490S, and 484Q. During the study, the 478K probe became available, and all samples with 452R were tested with this probe to evaluate for the delta variant. The N2RP assay was performed as described [20, 27]. For full Spike SNP methods, please see supplemental material.

### Sequencing

All rRT-PCR products from the first Spike SNP reaction (348-base-pair amplicon) were stored at 4°C and later shipped to GeneWiz (South Plainfield, NJ) for Sanger sequencing. Amplicon sequence was considered to be high quality and included for analysis if it achieved a quality score ≥ 40 and had base calls for all probe targets in at least one direction. A quality score ≥ 40, as defined by the company, signifies a mean probability of error ≤ 0.01% for each base call across the amplicon.

Complete genome sequencing was performed following the Artic Network protocol using the V3 primers scheme [28]. The genomic libraries were prepared and then loaded on a R9.4.1 flow cell (ONT FLO-MIN106) for sequencing with MinION device (MIN-101B). Basecalling, demultiplexing and trimming was carried out by Guppy toolkit integrated in the MinKNOW Nanopore software (Oxford Nanopore Technologies, UK). Genome assembly and consensus sequences were obtained with Nanopore EPI2ME (using Flye and Medaka respectively). The Nextclade tool was used for clade definition and mutation calling [29] and lineages were assigned using Pangolin web server [30]. Sequences were deposited in the GISAID database with accession numbers provided in Table 3.

### Statistics

Basic statistical analyses were performed using GraphPad Prism version 9.0.2 (GraphPad Software, San Diego, CA).

## Results

### Spike SNP optimization

For initial testing, a Spike SNP assay was developed that contained probes for 452Q, 484Q, and 490S. With the global emergence of the B.1.617.2 and lack of 484Q detection, the 478K probe was incorporated in the assay in place of 484Q. Addition of the 478K probe had no impact on 452Q or 490S detection (Figure 2), and despite overlapping sequences, the 478K and 484Q probes demonstrated no interference when combined in the same reaction (Figure S1). New probes for 484K and 501Y without LNA bases provided sensitive detection at both sites, though Ct values with the unmodified 484K probe were ∼2 cycles later than those generated with the LNA probe (Figure S2). Testing of clinical samples in the current study was performed with the original 484K and 501Y probes to maintain consistency with existing protocols. All primers and probes used in or developed for this study are shown in Table 1, and their respective locations across the amplicon are displayed in Figure 1.

**Table 1.**
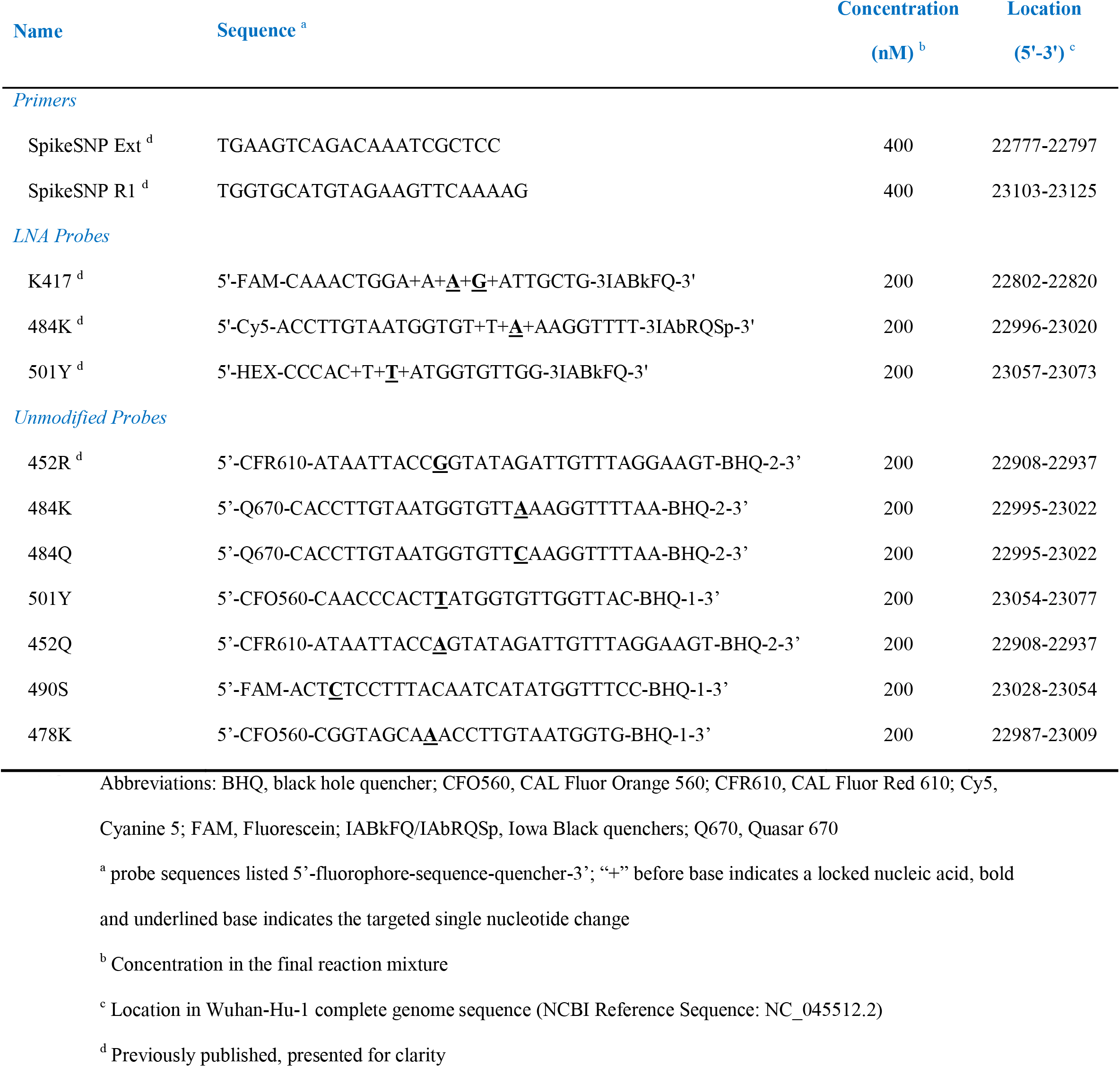
Spike SNP assay primers and probes used in the current study.

**Figure 2.**
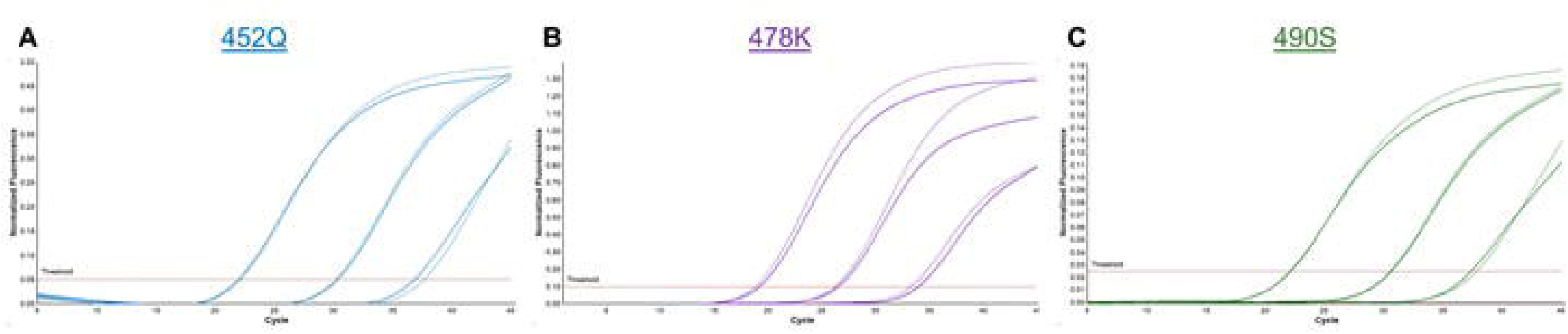
Addition of 478K probe does not impact signal from 452Q or 490S probes. Amplification curves are shown for 100-fold dilutions of C.37 (lambda) and B.1.617.2 (delta) samples tested together on a single run. Results were similar for **A**) 452Q and **C**) 490S mutations in C.37 with the probe for 478K (solid curves) or without (dotted curves). Results for B.1.617.2 were similar with the 478K probe (**B**) used in singleplex (dotted curves) or triplex reactions (solid curves). Nonspecific signals were not detected in any channel.

### SNP detection in clinical samples

Clinical samples were collected between November 2020 and April 2021. 113 participants were female (56.8%, of 199 with gender available), and age and day of symptoms at sample collection are shown in Table 2 along with N2RP and Spike SNP assay results. The distribution of N2 target Ct values is shown in Figure 3. The most common genotype identified was 417variant/484K/501Y (102/198 samples, 51.5%), which is consistent with B.1.351 (beta) and P.1. Large numbers of cases were also detected with signal for K417 only (64/198, 32.3%), consistent with a non-variant/ancestral lineage, and K417/484K (22/198, 11.1%), which is observed in P.2 (zeta), among others. Seven samples (3.5%) had the genotype K417/452R but tested negative with the 478K probe and were thus not consistent with B.1.617.2. Samples with the 417variant/484K/501Y genotype had significantly lower N2 Ct values (mean 19.3, standard deviation 5.0) than all other samples (22.0, 5.4; p < 0.001, data not shown) and samples with K417 only (22.1, 5.4; p = 0.002; Figure 3B). No differences in patient age or day of symptoms were observed based on genotype (p > 0.05 for all comparisons).

**Table 2.** Genotypes detected in the Spike SNP assay, patient demographics and the percent confirmed by amplicon sequencing.

| Category | n (%) | Age, years,<br>mean (SD) <sup>a</sup> | Day of Symptoms,<br>Mean (SD) <sup>b</sup> | Ct, mean (SD) <sup>c</sup> | Sequencing<br>Confirmed, n/N (%) <sup>d</sup> |
| --- | --- | --- | --- | --- | --- |
| N2 detected | 201 (100) | 42.6 (16.4) | 4.8 (3.0) | 20.8 (5.6) | — |
| Spike SNP detected | 198 (98.5) | 42.5 (16.4) | 4.8 (3.0) | 20.6 (5.3) | 181/181 (100) |
| *Genotype* <sup>e</sup> |  |  |  |  |  |
| 417var/484K/501Y | 102 (51.5) | 42.8 (14.9) | 4.5 (2.4) | 19.3 (5.0) | 96/96 (100) |
| K417 | 64 (32.3) | 42.6 (18.8) | 5.1 (3.7) | 22.1 (5.4) | 56/56 (100) |
| K417/484K | 22 (11.1) | 37.4 (14.2) | 4.8 (2.7) | 22.2 (5.4) | 20/20 (100) <sup>f</sup> |
| K417/452R | 7 (3.5) | 54.0 (15.8) | 5.0 (4.6) | 19.4 (4.0) | 7/7 (100) |
| K417/490S | 1 (0.5) | 41 | 4 | 15.2 | 1/1 (100) |
| K417/501Y | 1 (0.5) | 68 | 5 | 19.7 | 1/1 (100) |
| K417/484K/501Y | 1 (0.5) | 19 | 10 | 32.1 | NA |

Abbreviations: var, variant; SD, standard deviation
<sup>a</sup> Age available for 194/201 (96.5%) samples
<sup>b</sup> Day of symptoms available for 136/201 (67.7%) samples
<sup>c</sup> Ct value in the N2 assay
<sup>d</sup> Number with sequence confirmed (n) over the number of amplicons with high quality sequence and base calls at each SNP position (N)
<sup>e</sup> Displayed as % of Spike SNP positive samples
<sup>f</sup> One sample had a mixed base (A/G) at the position targeted by the 484K probe

**Figure 3.**
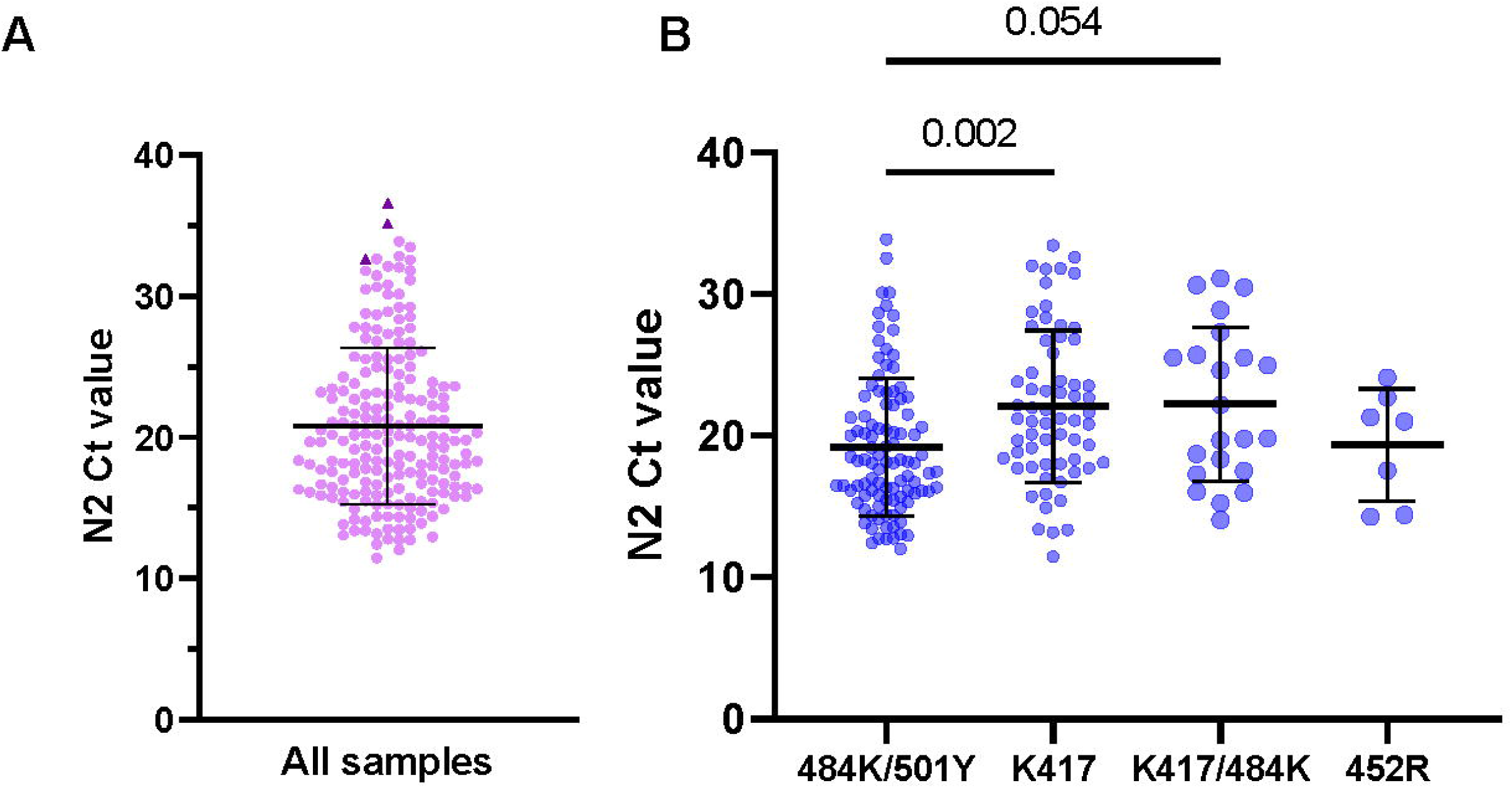
Distribution of N2 Ct values for A) all samples and B) the four most common genotypes detected. A) N2 Ct values for all tested samples are displayed. Three samples that tested negative in the Spike SNP assay are highlighted (dark triangles) and had N2 Ct values of 32.6, 36.6, and 35.1. B) N2 Ct values for the four most common genotypes detected. Bars represent mean Ct and standard deviation, and p-values for tested comparisons are displayed above the graph.

### Amplicon and whole genome sequencing

High quality amplicon sequences were obtained from 181 samples (181/201, 90.0%) by Sanger sequencing, and results confirmed Spike SNP genotype calls for all samples (Table 2). Characteristic findings for samples with 484K, with and without 501Y, are shown (Figure 4). All samples with variant calls at position 417 had the mutation AAG>ACG conferring K417T observed in P.1. One sample had 484K detected in the Spike SNP assay (genotype K417/484K) but E484 based on the consensus Sanger sequence. On review of the tracing, there was a mixture of G and A bases at the first position of codon 484, consistent with evidence of a mixed viral population in this sample. Notably, this sample had a late Ct value for 484K (38.4) relative to K417 (34.3) when compared to other samples with this genotype (mean Ct difference 0.4, standard deviation 0.2). In addition to mutations targeted in the Spike SNP assay, 4 samples (2.2%) had a mutation conferring T478R (ACA>AGA), and one sample (0.5%) had a mutation resulting in N501T (AAU>ACU).

**Figure 4.**
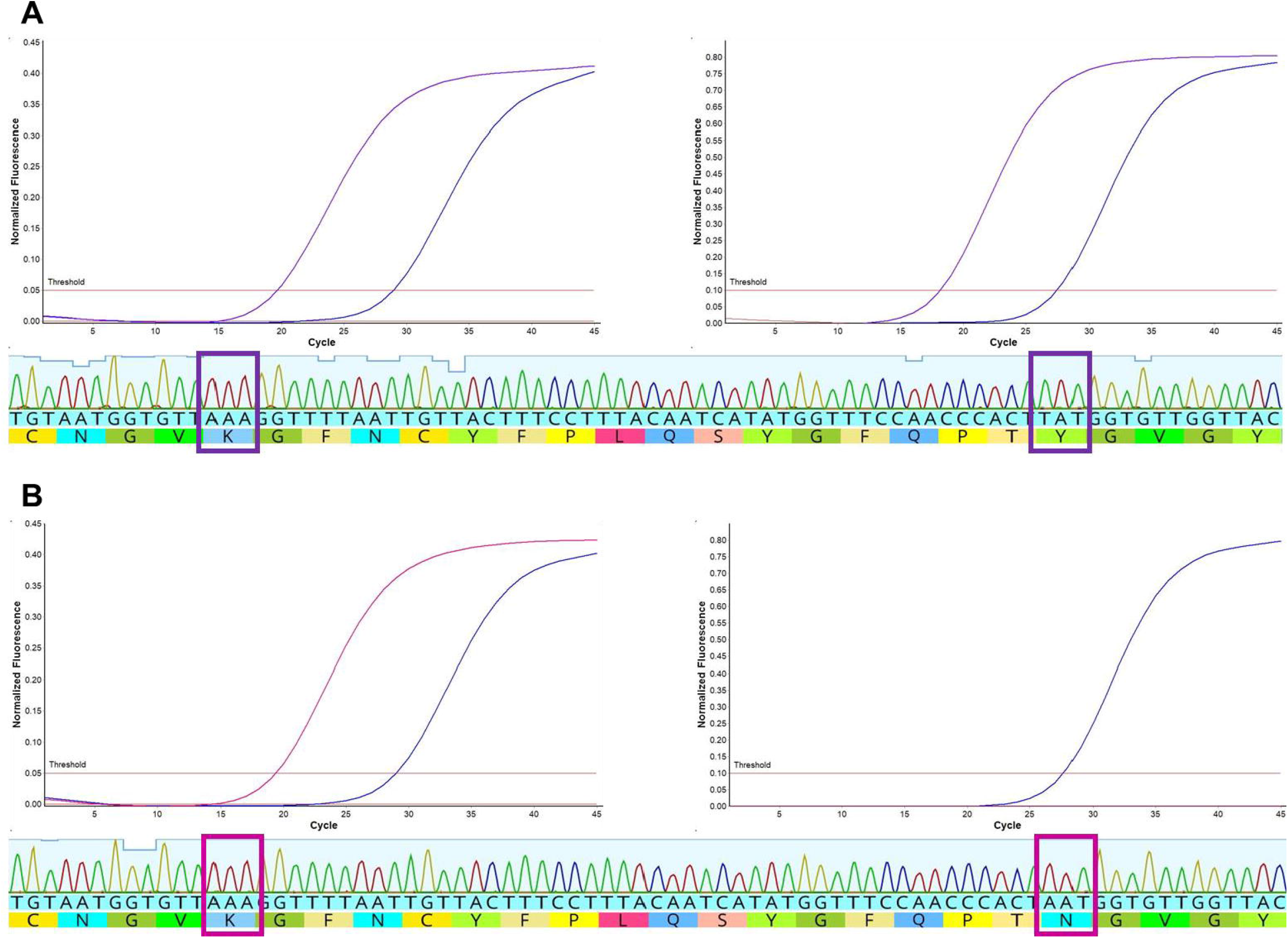
Spike SNP and amplicon sequencing results are concordant. Amplification curves and amplicon sequence tracings are shown for samples with mutations encoding **A**) 484K and 501Y (purple curves) or **B**) 484K only with N501 (cranberry). A beta variant control sample is shown for reference (blue curves).

Whole genome sequences for 29 samples were obtained using Artic protocol. Ten different lineages were identified (Table 3) [30]. Nanopore sequencing confirmed Spike SNP genotype calls for all samples. Samples bearing genotype 417variant/484K/501Y belonged to the P.1 lineage and P.1.2 sub-lineage. Genotype of only K417 was found in samples of lineages B.1.1.28, B.1.1.33, B.1.1.277 and N3 all within 20B Nextstrain clade and lineage B.1.499 (clade 20C) [29]. The P.2 lineage that also belongs to 20B clade showed the K417/484K genotype. One sample with genotype K417/501Y was classified as B.1.1.7 (alpha). A lineage from 2020 that was still circulating during 2021, A.2.5.2, was found in one sample with genotype K417/452R.

**Table 3.** Spike mutations and identified lineages by genome sequencing.

| Sample Code | Spike SNP Genotype | Nanopore Sequencing |  |  |  |
| --- | --- | --- | --- | --- | --- |
|  |  | GISAID Accession No. | Nextstrain Clade WHO label | Pango Lineage | Spike deletions /amino acid substitutions<sup>a</sup> |
| PY21-100 | 417var/484K/501Y | EPI_ISL_4071892 | 20J (V3) gamma | P.1 | L18F/T20N/P26S/D138Y/R190S/**K417T/E484K/N501Y**/D614G/ H655Y/T1027I/ V1176F |
| PY21-9 | 417var/484K/501Y | EPI_ISL_2444778 | 20J (V3) gamma | P.1 | L18F//T20N/P26S/D138Y/R190S/**K417T/E484K/N501Y**/D614G/ H655Y/T1027I/V1176F |
| PY21-57 | 417var/484K/501Y | EPI_ISL_4137476 | 20J (V3) gamma | P.1 | L18F/T20N/P26S/D138Y/R190S/**K417T/E484K/N501Y**/D614G/ H655Y/T1027I/V1176F |
| PY21-58 | 417var/484K/501Y | EPI_ISL_4071897 | 20J (V3) gamma | P.1 | L18F/T20N/P26S/D138Y/R190S/**K417T/E484K/N501Y**/D614G/ H655Y/T1027I/V1176F |
| PY21-84 | 417var/484K/501Y | EPI_ISL_4071898 | 20J (V3) gamma | P.1 | L18F/T20N/P26S/D138Y/R190S/**K417T/E484K/N501Y**/D614G/ H655Y/T1027I/V1176F |
| PY21-16 | 417var/484K/501Y | EPI_ISL_4071893 | 20J (V3) gamma | P.1 | L18F/T20N/P26S/D138Y/R190S/**K417T/E484K/N501Y**/D614G/ H655Y/T1027I/V1176F |
| PY21-11 | 417var/484K/501Y | EPI_ISL_4071900 | 20J (V3) gamma | P.1 | L18F/T20N/P26S/D138Y/L189F/R190S/**K417T/E484K/ N501Y**/ D614G/H655Y/T1027I/V1176F |
| PY21-90 | 417var/484K/501Y | EPI_ISL_4071899 | 20J (V3) gamma | P.1.2 | L18F/T20N/P26S/T95I/D138Y/R190S/**K417T/E484K/N501Y**/D614G/H655Y/ Q675H/T1027I/V1176F/V1228L |
| PY21-150 | K417 | EPI_ISL_2234899 | 20B | B.1.1.33 | D614G |
| PY21-153 | K417 | EPI_ISL_2234891 | 20B | B.1.1.33 | D614G |
| PY21-140 | K417 | EPI_ISL_2234880 | 20B | B.1.1.28 | D614G/V1176F |
| PY21-145 | K417 | EPI_ISL_4084605 | 20B | B.1.1.28 | D614G/V1176F |
| PY21-146 | K417 | EPI_ISL_2234881 | 20B | B.1.1.28 | D614G/V1176F |
| PY21-98 | K417 | EPI_ISL_2444788 | 20B | B.1.1.28 | D614G/V1176F |
| PY21-152 | K417 | EPI_ISL_2234885 | 20B | B.1.1.28 | T22I/D614G/V1176F |
| PY21-144 | K417 | EPI_ISL_4071901 | 20B | B.1.1.28 | D614G/N1135Y/V1176F |
| PY21-43 | K417 | EPI_ISL_4071895 | 20B | B.1.1.28 | A575S/D614G/V1176F |
| PY21-44 | K417 | EPI_ISL_4084604 | 20B | B.1.1.28 | T572I/D614G/V1176F |
| PY21-47 | K417 | EPI_ISL_4071894 | 20B | B.1.1.28 | A575S/D614G/V1176F |
| PY21-148 | K417 | EPI_ISL_2234894 | 20B | B.1.1.28 | V6F/L18F/D614G/V1176F |
| PY21-147 | K417 | EPI_ISL_2234887 | 20B | B.1.1.28 | Q14H/D614G/D1153A/V1176F |
| PY21-154 | K417 | EPI_ISL_2234893 | 20B | B.1.1.277 | D614G/V1176F |
| PY21-52 | K417/484K | EPI_ISL_2444780 | 20B (zeta) | P.2 | **E484K**/D614G/V1176F |
| PY21-157 | K417/484K | EPI_ISL_2234879 | 20B (zeta) | P.2 | L5F/**E484K**/D614G/A771S/V1176F |
| PY21-143 | K417 | EPI_ISL_2234889 | 20B | N.3 | T76I/D614G |
| PY21-141 | K417 | EPI_ISL_2234883 | 20C | B.1.499 | T478R/D614G |
| PY21-149 | K417 | EPI_ISL_2234897 | 20C | B.1.499 | T478R/D614G |
| PY21-102 | K417/452R | EPI_ISL_4071896 | 19B | A.2.5.2 | **L452R**/D614G |
| PY21-156 | K417/501Y | EPI_ISL_4084606 | 20I (V1) alpha | B.1.1.7 | DEL69-70/DEL144 **N501Y**/A570D/D614G/P681H/T716I/S982A/ D1118H |

Abbreviations: var, variant; WHO, World Health Organization
<sup>a</sup> Bold text indicates amino acid substitutions detected by the Spike SNP assay.

## Discussion

Among individuals with SARS-CoV-2 infection in Paraguay, the Spike SNP assay provided accurate detection of targeted mutations in the RBD and, in conjunction with the employed sequencing protocols, confirmed the predominance of B.1.1.28, P.2 and P.1 in this population between November 2020 and April 2021, as well as the relative absence of B.1.1.7. Given frequent travel between Paraguay and Brazil, B.1.1.28 lineage was expected to account for a high proportion of cases in late 2020 and early 2021, to be later replaced by P.1 and P.2. Samples with P.1 had lower Ct values in our population. A similar finding was documented initially in Brazil and likely results in increased transmissibility of gamma variant with replacement of other lineages [31, 32]. P.1 and P.2 lineages were readily differentiated from one another in the Spike SNP assay, and in a real-world scenario, N2RP and Spike SNP assays could be performed sequentially to diagnose SARS-CoV-2 infection, identify a set of common mutations, and select samples for downstream sequencing [4, 5, 20].

For the current study, new probes were generated for mutations that occur within the target region and are associated with emerging VOCs/VOIs, including delta and lambda variants (Figure 1). As new designs do not involve separate primer-probe sets, the assay can be readily modified and reconfigured to include probes that match circulating variants in a particular region. All new probes were designed without LNA bases yet demonstrated similar performance to previous designs (Figure S1). Although LNA bases have proven important for detection of K417 mutations, avoiding their use in other probes improves access to and reduces costs for these oligos. An additional benefit to the current design is that up to 4 mutations are targeted in a single reaction, which has been found to improve variant classification [9]. Other published rRT-PCR protocols for the detection of a similar number of mutations generate 2 or more amplicons in single-reaction [12, 15, 17] or multiple-reaction designs [6, 8, 14, 16], and such protocols may be more difficult to modify in response to the emergence of new variants.

Sequencing methods employed as part of our testing protocol included whole genome sequencing on a MinION device and Sanger sequencing of the Spike SNP amplicon. Nanopore sequencing has become a common method worldwide and particularly in locations where use of short-read sequencing platforms is less feasible. MinION sequencing has been well described for SARS-CoV-2 and provides formal confirmation of detected variants, which cannot be achieved by rRT-PCR alone [4, 5, 25, 31]. Amplicon sequencing provides less information, but as demonstrated in this study, it serves to confirm Spike SNP calls, differentiate B.1.351 and P.1, and identify additional mutations in the receptor binding motif. In our study, 4 individuals had the T478R mutation; the whole genome of two of them were sequenced and were assigned the B.1.499 lineage. This lineage circulated widely in Argentina during 2020, but the mutation T478R was not reported. While this mutation is distinct from T478K found in the B.1.617.2 (delta), mutations at position 478 have developed in vitro and confer decreased antibody neutralization, indicating that such mutations may benefit the virus through immune escape [33, 34]. Finally, one individual was identified with a possible mixed infection or a minor variant bearing 484K [35, 36]. Such infections do not appear to cause more severe clinical disease [35], but this case highlights one potential benefit of rRT-PCR surveillance if key mutations can be detected that are not observed in the consensus sequence.

The degree of multiplexing possible in the Spike SNP assay is limited by the number of channels available in common real-time PCR instruments (4-5) as well as the defined 348-base-pair target region. Based on results of testing for 478K and 484Q in a single reaction, overlapping probes do not impact performance (Figure S1). However, multiplexed detection of key mutations outside of the Spike SNP target region would require additional primer-probe sets and assay optimization. A limitation to the current study is the utilization of retrospective convenience samples to evaluate the testing protocol. Although new data on SARS-CoV-2 variants in Paraguay were generated, we cannot calculate variant prevalence from these data.

In conclusion, the Spike SNP assay provides accurate detection of mutations that are associated with VOCs/VOIs in Paraguay. This can be implemented in SARS-CoV-2 testing protocols to triage samples for whole genome sequencing in the MinION platform or direct amplicon sequencing, which provides additional information. The assay can be implemented in any laboratory performing rRT-PCR to improve surveillance for these mutations and inform the judicious use of scarce sequencing resources.

## Data Availability

Data will be made available upon request following acceptance of this manuscript for publication.

## Acknowledgements

The authors thank the staff at the Instituto de Investigaciones en Ciencias de la Salud, Universidad Nacional de Asunción, and collaborating laboratories: Letizia Carpinelli (CYRLAB), Fátima Ovando (Hospital de Clínicas, UNA), Graciela Riera (MeyerLab) and Stefan Goertzen (Hospital Loma Plata) for their support during the study period and the COVID pandemic in Paraguay. We also thank members of the Variant Task Force at Emory University, Aunradha Rao, Leda Bassit, and Morgan Greenleaf, for the provision of samples with characterized variant strains for assay evaluation.

## Funding

This work was partially funded by Consejo Nacional de Ciencia y Tecnología with FEEI support, CONACYT, Paraguay; PINV20-239 (MM) and the Doris Duke Charitable Foundation, Clinical Scientist Development Award 2019089478 (JJW)

## Conflict of Interests

Authors declare that they have no conflict of interest.

## Notes

### Competing Interest Statement

The authors have declared no competing interest.

### Funding Statement

This work was partially funded by Consejo Nacional de Ciencia y Tecnologia with FEEI support, CONACYT, Paraguay; PINV20-239 (MM) and the Doris Duke Charitable Foundation, Clinical Scientist Development Award 2019089478 (JJW)

### Author Declarations

This study was reviewed and approved by the IICS Scientific and Ethics Committee (P38/2020) and the Emory Institutional Review Board (study 00110736).

