## Supplemental Material for "SARS-CoV-2 variants in Paraguay: Detection and surveillance with a readily modifiable, multiplex real-time RT-PCR"

Magaly Martinez and Phuong-Vi Nguyen, Maxwell Su, Fátima Cardozo, Adriana Valenzuela, Laura Franco, María Eugenia Galeano, Leticia Elizabeth Rojas, Chyntia Carolina Díaz Acosta, Jonás Fernández, Joel Ortiz, Florencia del Puerto, Laura Mendoza, Eva Nara, Alejandra Rojas and Jesse J. Waggoner

### Supplemental Methods

#### Spike SNP Performance

Spike SNP assays were performed as separate runs on the same day using the following conditions: 52°C for 15 min, 94°C for 2 min, and 45 cycles of 94°C for 15 sec and 60°C for 60 sec. At 60°C, fluorescent signal was acquired in all channels. Thresholds for each channel were set during assay optimization and used for all subsequent analyses. During analysis of the Spike SNP data, outlier removal was performed for signals that did not reach 10% (K417, 490S) or 20% (452R, 484K, 501Y, 452Q, 484Q, 478K) of the maximum fluorescence in a given channel.

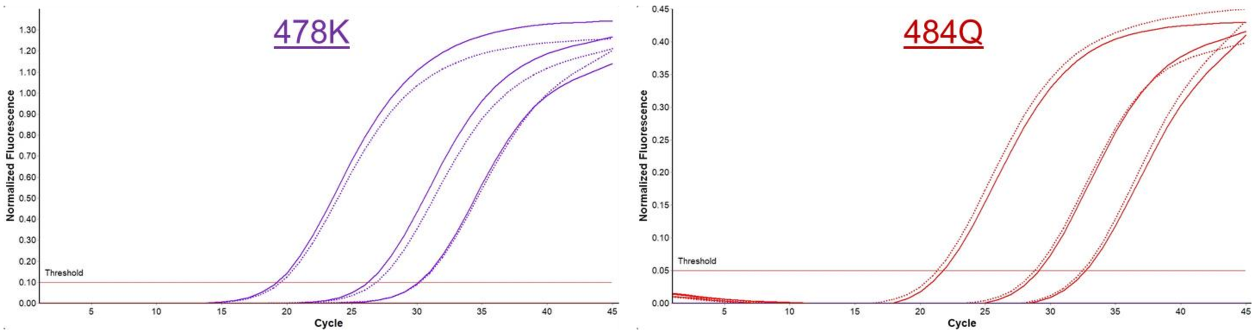


**Figure S1. 478K and 484Q probes do not interfere with detection despite overlapping sequences.** Dilutions of RNA from B.1.617.2 (delta variant, 478K) and B.1.617.1 (484Q) were tested in Spike SNP triplexes containing probes for 452Q, 490S, and either 478K or 484Q (solid curves), or a Spike SNP quadruplex assay containing all 4 probes on a single run (dotted curves). Amplification curves are shown for undiluted eluate, 100-fold and 1000-fold dilutions for each variant.

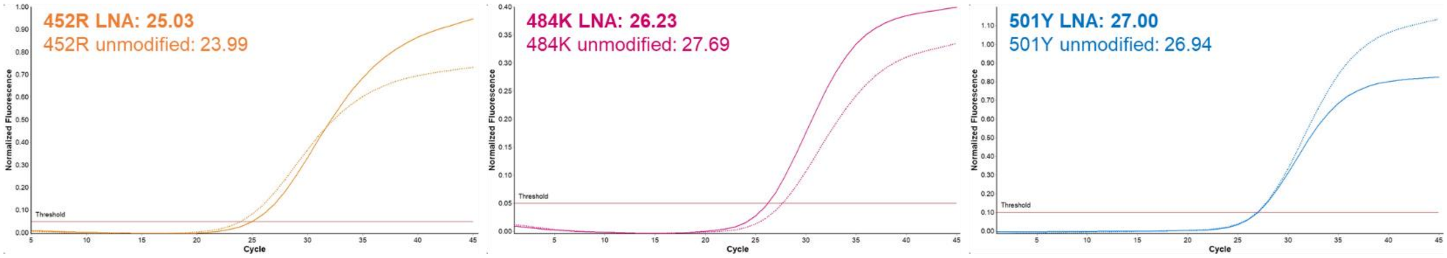


**Figure S2.** Performance of unmodified hydrolysis probes compared to probes that include LNA bases for the detection of mutations that cause 452R, 484K and 501Y. Controls containing the particular mutation were tested on a single run using both probes. Ct values are displayed. Curves generated from probes containing LNA bases are shown as solid lines; those generated with unmodified probes are displayed as thin, dotted lines.
